# Working conditions, security and grievance management among Bundibugyo virus response personnel: exploratory findings from an anonymous survey in eastern Democratic Republic of the Congo

**DOI:** 10.64898/2026.09.08.26362489

**Authors:** Johan G. L. Verheyden, Celestin Nzanzu Mudogo

**Affiliations:** Aries Consult Kenya, Nairobi, Kenya; Department of Basic Sciences, Unit of Molecular Biology and Biochemistry, School of Medicine, University of Kinshasa, Kinshasa, Democratic Republic of the Congo

**Author notes:** Corresponding author: Johan G. L. Verheyden —, Sepal Gardens, Apt 605, Gatunda Close, Kileleshwa, Nairobi, Kenya. Co-corresponding author: Celestin Nzanzu Mudogo —.

**Keywords:** Bundibugyo virus disease, Ebola, health workers, working conditions, grievances, security, infection prevention and control, Democratic Republic of the Congo

## Abstract

**Problem:** Response personnel in the 2026 Bundibugyo virus disease (BVD) outbreak in eastern Democratic Republic of the Congo work amid protracted conflict and fragile health systems. Working conditions, security exposure and grievance handling were widely reported as sources of stress but had not been documented systematically.

**Approach:** We conducted an anonymous, self-administered online survey in French among BVD response personnel in North Kivu, Ituri and South Kivu, covering staffing, EPI supply, remuneration, security incidents, grievances and psychosocial well-being. Of 55 submissions, 54 respondents consented and were analysed; denominators fall further, to 21, for grievance items because of within-survey drop-off. Findings are descriptive.

**Local setting:** Respondents (94% men) worked for the Ministry of Health, international non-governmental organizations, the Red Cross and as unpaid volunteers, mostly with one to three months on the response.

**Relevant changes:** 61% judged staffing insufficient; 67% had one or no rest day per week. 63% reported EPI stock-outs and 43% had performed a task judged riskier than necessary. 44% had pay delayed, partial or unpaid though promised, and 85% had advanced personal funds for work costs. 67% reported a security incident in three months; among the smaller subgroup reaching the grievance questions, 57% feared reprisal.

**Lessons learnt:** Staffing, supply, payment and security shortfalls compound one another and are visible to communities; weak grievance channels and fear of reprisal let them persist unaddressed. Working conditions, security and grievance management should be monitored as core infection-prevention-and-control and occupational-risk indicators, not as a peripheral human-resources matter.

**What was already known on this topic:** Previous Ebola epidemics, notably the 2014-2016 West African outbreak, showed that health-worker pay disputes, personal protective equipment shortages and community mistrust can contribute to absenteeism, disengagement and reduced service continuity [2,5,6], and that health workers’ coping depends on training, equipment, a functioning risk allowance and psychosocial support [2]. In eastern Democratic Republic of the Congo, repeated Ebola-family outbreaks have occurred alongside armed conflict and institutional distrust [1], but working conditions, security exposure and grievance handling among response personnel are rarely documented systematically during an active response.

**What this paper adds:** Using an anonymous survey administered during an ongoing 2026 BVD response, this paper quantifies, in near real time, how response personnel across three eastern DRC provinces experienced staffing, EPI supply, remuneration, security and grievance handling, and how these problems interact. It shows that administrative and security shortfalls are interconnected and visible to communities, and that survey drop-off is itself informative: fewer respondents were willing or able to complete questions on grievances and psychosocial strain than on operational conditions.

## Introduction

Epidemic response in complex humanitarian settings depends not only on epidemiological and clinical capacity but on a workforce that is adequately staffed, equipped, paid and protected. In eastern Democratic Republic of the Congo (DRC), responders operate amid protracted conflict, displacement, fragile health systems and recurrent viral haemorrhagic fever outbreaks. During the 2026 Bundibugyo virus disease (BVD) epidemic, personnel were deployed under diverse contractual arrangements across North Kivu, Ituri and South Kivu [7,8]. Reports of unpaid allowances, equipment shortages and insecurity circulated informally but had not been documented systematically. We implemented an anonymous survey to describe response personnel’s working conditions, security exposure and grievances, and to identify patterns with operational implications for infection prevention and control (IPC) and occupational risk.

## Methods

We designed a structured questionnaire covering role and employer; staffing, workload and rest; availability of personal protective equipment (EPI); remuneration, allowances and personal advances; security incidents and perceived risk; community relations; grievances and fear of reprisal; and psychosocial well-being. The survey was administered anonymously in French through an online platform during the response. Participation was voluntary; no directly identifiable information was collected. Of 55 submissions received, one respondent did not consent and was excluded, leaving 54 analysed responses. Not every respondent answered every item, and non-response increased through the questionnaire: core operational items (staffing, EPI, pay, security) were answered by all 54 respondents, but items on formal grievance mechanisms and psychosocial well-being, positioned later in the questionnaire, were reached by only 21. We report descriptive counts and proportions among respondents to each item, with denominators stated throughout, and present selected open responses, translated from French, to illustrate key themes. We applied no statistical inference; findings are exploratory signals from a small, self-selected sample.

## Results

### Respondent profile

Fifty-four respondents were analysed: 51 (94%) men and 3 (6%) women, mostly aged 25-44 years (36/54, 67%). Employers included the Ministry of Health (16/54, 30%), international non-governmental organizations (15/54, 28%), unpaid volunteer arrangements (15/54, 28%), and the Red Cross or local community organizations (3/54, 6%). Roles included physicians (13), community mobilisers/risk-communication staff (11), surveillance/investigation officers (8), infection-prevention-and-control/WASH and laboratory staff (5 each) and nurses (4), among others. Most respondents worked in North Kivu (23/54, 43%) or Ituri (19/54, 35%); most had one to three months on the current response (35/54, 65%).

### Staffing, workload and EPI supply

Sixty-one per cent of respondents (33/54) judged available staffing insufficient or very insufficient for the workload, against 15% (8/54) who judged it sufficient. Rest was limited: 31% (17/54) reported no rest day and 35% (19/54) one day per week. EPI availability was inconsistent: 63% (34/54) reported stock-outs at least sometimes, most often of protective suits (27/41, 66%), disinfectant or chlorine (24/41, 59%) and gloves (23/41, 56%). Forty-three per cent (23/54) had performed a task in the past three months that they judged riskier than necessary, most commonly working without adequate EPI despite a known risk (17/23, 74%) or operating in an area judged unsafe (13/23, 57%). One infection-prevention-and-control/WASH respondent summarised that a response is impossible without well-equipped active case-finding, a triage service to protect staff, and functioning infection prevention and control (Table 1; Fig. 1).

**Table 1.** Selected indicators from an anonymous survey of Bundibugyo virus response personnel, eastern Democratic Republic of the Congo, 2026 (N = 54 unless noted). *Subset of 21 respondents who reached the grievance section of the questionnaire.

| Indicator | n/N | % |
| --- | --- | --- |
| Judged available staffing insufficient or very insufficient | 33/54 | 61% |
| ≤1 rest day per week | 36/54 | 67% |
| EPI (personal protective equipment) reported out of stock at least sometimes | 34/54 | 63% |
| Risk-premium eligibility criteria unclear or not at all clear | 40/54 | 74% |
| Pay or allowance delayed, partial, or unpaid though promised | 24/54 | 44% |
| Advanced personal funds for work costs (at least rarely) | 46/54 | 85% |
| Performed a task judged riskier than necessary (past 3 months) | 23/54 | 43% |
| Felt free to refuse a risky task without negative consequence | 22/54 | 41% |
| Continued a risky task specifically because could not afford to lose the income | 24/54 | 44% |
| Rated insecurity in work area as moderate or higher | 41/54 | 76% |
| Experienced ≥1 security-related incident (past 3 months) | 36/54 | 67% |
| Low or very low confidence in organizational security measures | 24/54 | 44% |
| Aware of a formal grievance mechanism* | 12/21 | 57% |
| Moderate-to-very-high fear of reprisal for raising a grievance* | 12/21 | 57% |

**Fig. 1.**
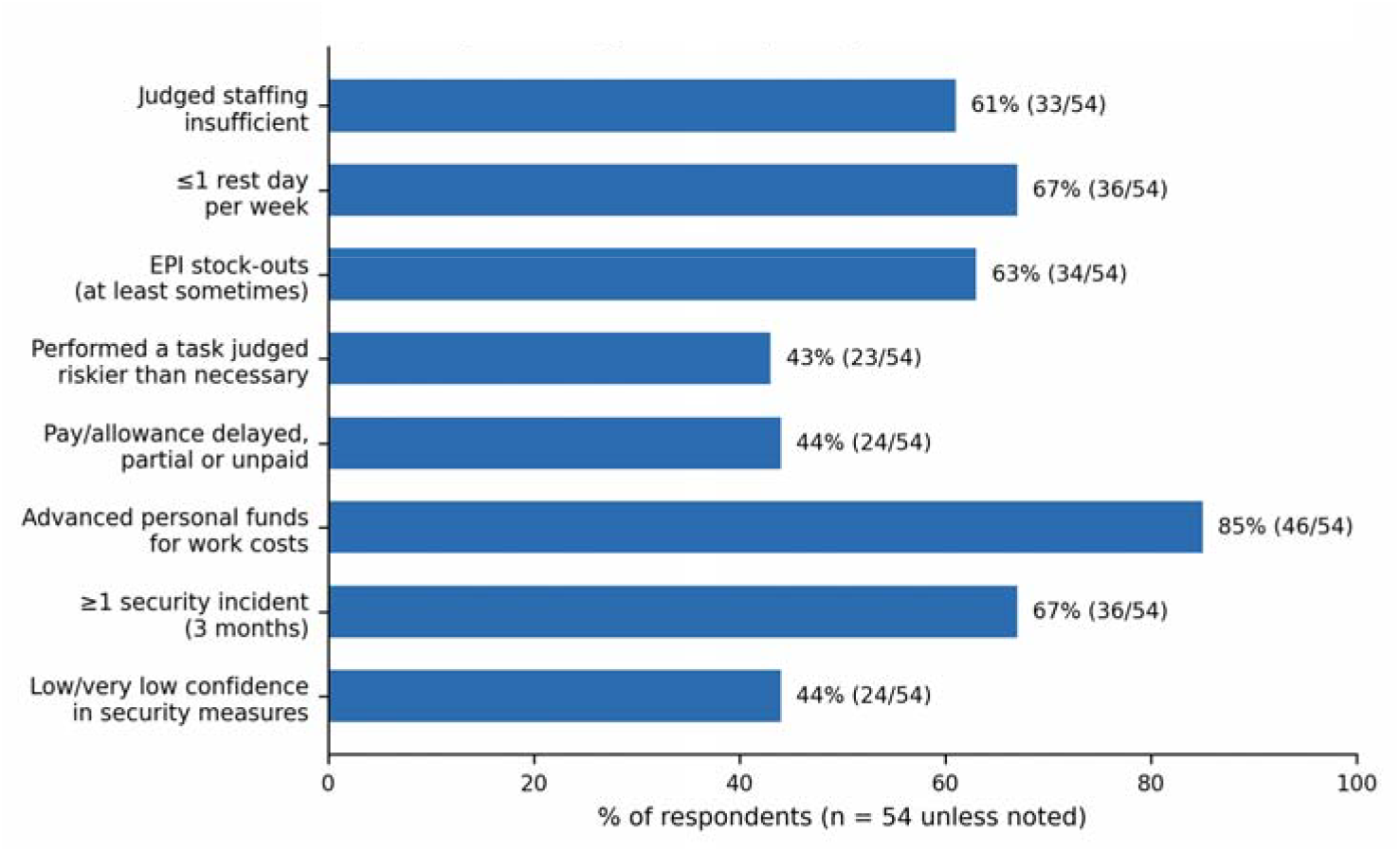
Selected working-condition, security and payment indicators reported by Bundibugyo virus response personnel, eastern Democratic Republic of the Congo, 2026 (n = 54).

### Remuneration, allowances and personal advances

Criteria for the risk allowance were unclear or not at all clear to 74% of respondents (40/54), and only 15% (8/54) had received it on time. Forty-four per cent (24/54) reported that pay or allowances promised to them had been delayed, only partially paid, or not paid at all; reported delays ranged from two weeks to more than a month. Eighty-five per cent (46/54) had advanced personal funds for work costs at least occasionally, most often for transport or fuel (35/46, 76%), communication (36/46, 78%) or food and water (33/46, 72%). A logistics respondent described teams buying their own fuel without being reimbursed, and another linked publicised donor funding to local frustration, since public figures for response funding circulated while personnel went unpaid. Perceived effects of payment problems, among 54 respondents with multiple answers permitted, included demotivation (24/54, 44%), indebtedness (19/54, 35%), interrupted activities (15/54, 28%) and, for 9/54 (17%), a strike, sit-in or other collective action.

### Security, refusal and constrained choice

Seventy-six per cent (41/54) rated insecurity in their work area as moderate or higher, and 67% (36/54) reported at least one security-related incident in the past three months, most often community hostility or aggression (21/54, 39%) or an armed-group threat (19/54, 35%); one respondent reported sexual or gender-based violence. Sixty-five per cent (35/54) had no secure transport for work travel, and 44% (24/54) reported low or very low confidence in their organization’s security measures. When a task was judged too risky, only 41% (22/54) felt free to refuse it without negative consequence, and 44% (24/54) said they had continued a risky task specifically because they could not afford to lose the income. One respondent described overriding an instruction to travel toward an area with reported unidentified armed elements by substituting remote data collection through community relays and phone-based reporting, then requesting a security corridor from logistics coordination, illustrating that risk-avoidance currently depends on individual initiative rather than standing policy.

### Grievances, fear of reprisal and psychosocial strain

Item non-response rose sharply for this section: only 21 of 54 respondents reached the grievance and psychosocial questions. Among them, 57% (12/21) were aware of a formal grievance mechanism, but only a quarter of those (3/12) expressed high confidence that a complaint would be acted on. Unresolved grievances most often concerned unpaid or delayed salary (12/21, 57%) and unmet commitments (7/21, 33%). Fear of reprisal for raising a grievance was moderate to very high for 57% (12/21); one respondent described threats against staff who declined to surrender a share of their risk allowance. Sixty-two per cent (13/21) reported feeling emotionally exhausted by their work at least sometimes in the past month, and 76% (16/21) had no access to psychosocial support.

## Discussion

These exploratory findings describe a self-reinforcing pattern: understaffing and long hours increase fatigue; EPI stock-outs and unclear entitlements push personnel toward personally risky choices or personal spending; delayed or opaque payments erode trust and motivation, feeding absenteeism and, occasionally, collective action; and security risk is compounded when personnel cannot refuse a task without financial consequence. Communities were reported to notice unpaid, under-equipped and absent personnel, which plausibly reinforces the mistrust and rumour dynamics already documented in this and earlier eastern DRC Ebola-family outbreaks. Grievances accumulate, but fear of reprisal and weak feedback loops appear to constrain corrective action, consistent with qualitative evidence that a functioning risk allowance, reliable supply and psychosocial support were central to how health workers coped during the 2014-2016 West African epidemic [2], and with evidence that institutional trust is fragile and consequential in this same setting [1]. WHO guidance already frames occupational risk management and infection prevention and control as linked domains [3,4]; these findings suggest that working conditions and grievance handling belong in the same frame.

Several limitations warrant caution. The sample is small (54 analysed responses) and self-selected, which may bias findings toward more, or less, dissatisfied personnel; sensitive topics may be under-reported even in an anonymous survey; and non-response rose sharply for grievance and psychosocial items, so those estimates rest on a smaller, potentially non-representative subgroup. No statistical inference was applied. These findings should be read as exploratory signals warranting operational follow-up, not as representative estimates for the full response workforce.

### Box 1. Lessons learnt and recommended actions

- Chronic understaffing and unclear rest patterns coexisted with EPI stock-outs, so tracking one indicator without the other risks missing their joint effect on IPC risk. A minimum operating package (functioning EPI, fuel/transport, communications, water, and a basic evacuation plan) should be verified for each active team.
- Delayed, partial or opaque payments were common (44% of respondents) and were linked by respondents to demotivation, indebtedness and, in some cases, collective action. The challenge is coordinating remittance across multiple employers (Ministry of Health, NGOs, Red Cross, volunteers); a shared, published payment schedule with named focal points per employer, checked against validated lists, is a low-cost first step.
- Personnel frequently could not refuse a risky task without financial or professional consequence — the same underlying income insecurity as the payment problem. Formalizing a no-penalty right to pause or decline a documented, dangerously risky task, alongside a mandatory pre-deployment risk assessment, would remove the current dependence on individual initiative illustrated in this survey.
- Fear of reprisal was common among the smaller subgroup who reached the grievance questions, and few trusted that a complaint would be acted on. A confidential, actively publicised grievance channel is needed — not only a policy on paper — since awareness of a mechanism did not translate into confidence in it.
- Non-response rose sharply for grievance and psychosocial items; this pattern is itself an operational signal that sensitive topics remain hard to surface even in an anonymous survey, and argues for triangulating survey data with confidential debriefs rather than treating a non-response as a null finding.
- Respondents reported that communities notice unpaid, under-equipped or absent personnel. Monitoring working conditions alongside community-engagement indicators, rather than as a separate human-resources track, would better reflect how these dynamics interact in the field.

## Data Availability

All data produced in the present study are available upon reasonable request to the authors

## Declarations

### Ethics

The survey was anonymous, voluntary and collected no directly identifiable information. It was designed as a minimal-risk exploratory instrument; ethics committee approval was not required under local practice, and findings are interpreted cautiously as exploratory signals.

### Data availability

The anonymized response-level dataset is available from the corresponding author on reasonable request. Given the small sample (n=54, falling to n=21 for grievance items), some response combinations could be identifying; requests will be reviewed to protect respondent anonymity before any data are shared.

### Author contributions

J.G.L.V.: conceptualization, survey design, data curation, formal analysis, writing — original draft. C.N.M.: epidemiological and clinical interpretation, validation, writing — review and editing. Both authors approved the final manuscript and, per ICMJE criteria, take public responsibility for its content.

### Declaration of AI assistance

During manuscript preparation, the authors used Claude (Anthropic; Claude Sonnet 5) under supervision to assist with data tabulation, organisation, language polishing and formatting. All analytic definitions, numerical results, interpretations and final text were reviewed and accepted by the authors, who take full responsibility for the accuracy and integrity of the work.

